# aiHumanoid Simulations Uncover Dominant-Negative Effects in HNRNPH2-Related Neurodevelopmental Disorders

**DOI:** 10.1101/2024.08.21.24312358

**Authors:** WR Danter

## Abstract

HNRNPH2-related neurodevelopmental disorders (NDDs) encompass a spectrum of cognitive, motor, and systemic impairments driven by mutations in the HNRNPH2 gene. While these disorders have been linked to both gain-of-function (GOF) and loss-of-function (LOF) mutations, the precise mechanisms underlying their varied phenotypic features remain unclear. In this study, we employed advanced aiHumanoid simulations to conduct a virtual longitudinal analysis of HNRNPH2 mutations, spanning from birth through young adulthood. Our findings reveal that LOF mutations, particularly severe missense, and nonsense/frameshift mutations, exert dominant-negative effects that significantly impair neurodevelopmental outcomes.

Through detailed comparison of mild versus severe mutations, we observed that severe LOF mutations lead to markedly worse cognitive, motor, and systemic deficits. These effects are compounded by the interference of mutant HNRNPH2 with the function of wild-type HNRNPH2 and potentially compensatory proteins such as HNRNPH1. Notably, our simulations indicate that the upregulation of HNRNPH1 observed in knockout models fails to compensate in the presence of dominant-negative mutations, highlighting a critical pathway disruption.

This study not only elucidates the mechanistic basis of dominant-negative effects in HNRNPH2-related NDDs but also suggests new therapeutic avenues, including the enhancement of compensatory mechanisms and targeted inhibition of mutant HNRNPH2. The use of aiHumanoid simulations offers a powerful tool for predicting long-term outcomes and testing potential interventions, providing a robust platform for advancing the understanding and treatment of HNRNPH2-NDDs.

Our findings underscore the importance of early diagnosis and intervention, particularly for patients with severe LOF mutations, and facilitate future research focused on mitigating the dominant-negative effects of these mutations.

## Introduction

Neurodevelopmental disorders (NDDs) associated with mutations in the HNRNPH2 gene [1,2] are characterized by a spectrum of developmental delays and impairments in cognitive, social, behavioral, and emotional functioning. Traditional models have often provided limited insights into the complex and dynamic processes involved in neurodevelopment. However, recent advancements in humanized brain simulation technologies have offered new ways to explore these disorders with greater accuracy and relevance to human brain development.

The use of cerebral organoids for modeling human brain development was first demonstrated by Lancaster et al. [3], who showcased their usefulness for studying conditions like microcephaly. This groundbreaking work laid the foundation for using organoid models to investigate a broader range of neurological conditions. Pasca [4] further highlighted the transformative impact of three-dimensional human brain cultures, emphasizing their ability to reproduce the structural and functional complexities of the human brain in vitro. Wang [5] expanded on this by exploring the construction of complex human brain models to deepen our understanding of both disease and normal development.

In the specific setting of genetic neurodevelopmental disorders, researchers such as Korff et al. [6] and Taylor et al. [7] have made significant contributions by elucidating the pathological mechanisms linked to HNRNPH2 mutations. These studies emphasize the critical role that genetic factors play in NDDs and point to potential therapeutic targets within these pathways.

Building on these foundational studies, artificially induced Whole Brain Organoid (aiWBO) models were developed to simulate complex diseases, such as Alzheimer’s and metachromatic leukodystrophy (MLD). Esmail and Danter [8,9] successfully utilized these aiWBO models, demonstrating their potential for disease modeling. More recently, advancements that involve combining multiple organoid simulations have led to the development of aiHumanoid simulations [10,11,12], which offer several advantages over aiWBO models, including more sophisticated representations of human brain physiology in the context of a simulated human and the ability to conduct early-stage virtual drug trials.

This paper aims to leverage the advanced capabilities of aiHumanoid simulations to conduct a virtual longitudinal study of HNRNPH2-related NDD in children and young adults. By integrating genetic, cellular, and molecular data into these simulations, we seek to unravel the functional impacts of HNRNPH2 mutations on brain development and function. The use of aiHumanoid simulations will allow us to predict the phenotypic outcomes of various HNRNPH2 mutations and identify potential therapeutic interventions over time. This approach not only promises to enhance our understanding of the development and progression of HNRNPH2-NDD but also sets the stage for future virtual drug trials, potentially revolutionizing treatment strategies for this rare group of genetic disorders.

## METHODS

### Overview

This project utilizes aiHumanoid simulations of 25 virtual subjects in a longitudinal study of the development and progression of HNRNPH2-associated Neurodevelopmental disorder (NDD) from birth to age 20 years. The primary objective is to better understand the systemic impact of HNRNPH2 mutations.

In this first phase of our HNRNPH2-NDD project, we will focus on the impact of mild and severe HNRNPH2 mutations on systemic and nervous system markers and outcomes. We anticipate that future research will build on this approach to develop early stage virtual drug trials aimed at identifying phenotype modifying therapies for HNRNPH2 related and other NDDs.

#### 1. Updating the aiHumanoid Simulation to v8.4.3

The previous version 8.4.2 of the aiHumanoid [12] underwent revisions to v8.4.3. The main differences are that the revised version integrates updated simulations for specific HNRNPH1 and NHRNPH2 associated mutations and an updated subsystem for the diagnosis of HNRNPH2-NDD in children and adolescents. As before, the number of integrated organoid simulations remains at 21. The literature validation of the WT and HNRNPH2 aiHumanoid simulations employed the same approach used in previous versions to create the updated simulations comprising v8.4.3. [12]

#### 2. Validation of the HNRNPH2-NDD Feature Profile in the aiHumanoid Simulations

To confirm a diagnosis of HNRNPH2-NDD in the affected aiHumanoid simulated young subjects, a list of seventeen genotypic and phenotypic features, with references, was assembled from the peer reviewed literature for evaluation and are presented in Appendix A. All features were statistically significantly different from controls for multiple age matched cohorts and regarding the specific mutation. The present statistical analysis employed a combination of the nonparametric Wilcoxon signed rank test and the Cliff’s delta effect size estimates.

#### 3. Study Design and Objectives

This project is our most recent virtual longitudinal study using the aiHumanoid simulations. The objectives of this study were: (i) to evaluate the impact of specific GOF and LOF mutations on young subjects compared to WT/Healthy subjects, (ii) to better understand HNRNPH2-NDD development and progression from birth to age 20 years, and (iii) to evaluate a panel of disorder features for the purpose of conducting future virtual drug trials to identify potential phenotype modifying therapies.

### The Virtual subjects used in this study

The profiles for twenty-five unique and healthy virtual young subjects were synthesized by GPT-4, an advanced large language model (December 2023 version,) at https://chat.openai.com/). GPT4 used its extensive database encompassing medical literature, patient profiles, and related clinical information, to synthesize diverse and representative draft profiles for twenty-five healthy children. Each subject profile was reviewed by an experienced physician prior to enrolment. This longitudinal study design permitted us to create a virtual study with 6 genotypic cohorts (see Table 1) X 6 (age groups) X 25 (virtual subjects), the equivalent of data from 900 young subjects. The virtual subjects used in this study are hypothetical, but commonly encountered population based examples of risks associated with the development of NDD in children in specific affected cohorts but do not represent actual individuals or precise medical histories.

**Table 1:**
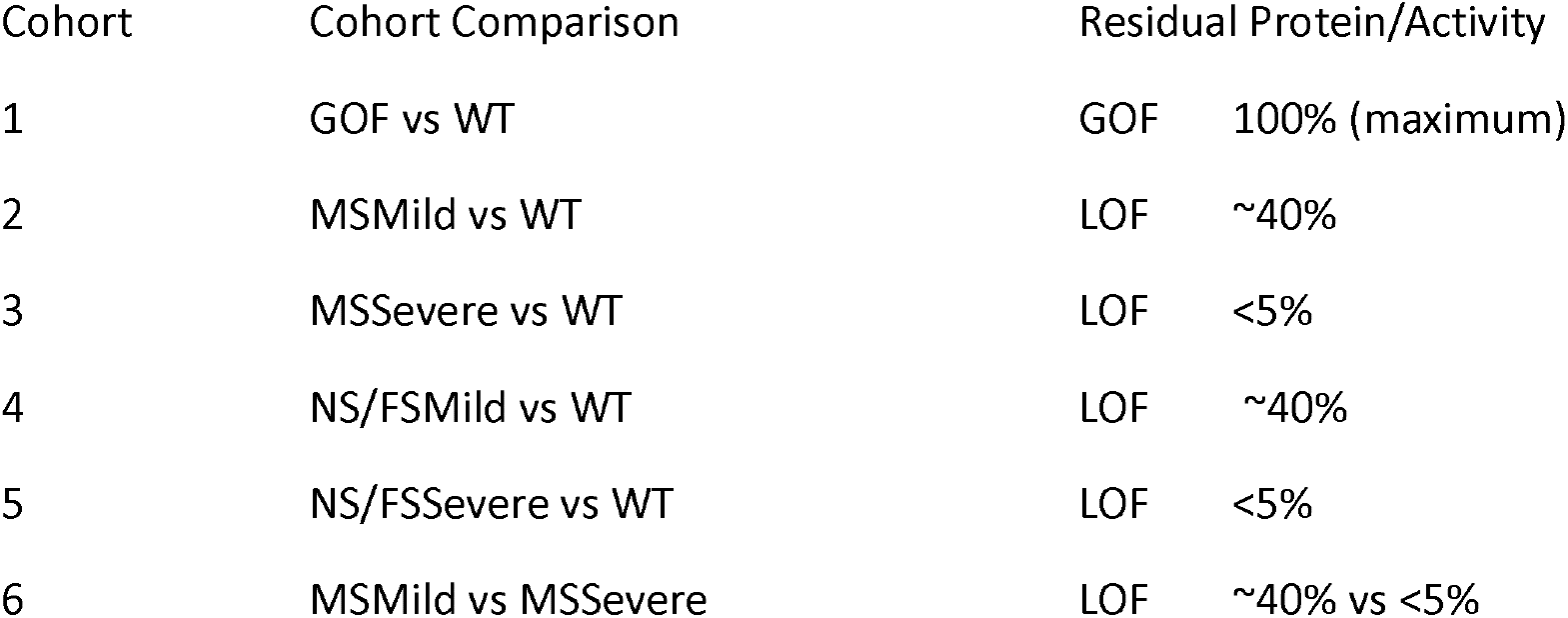
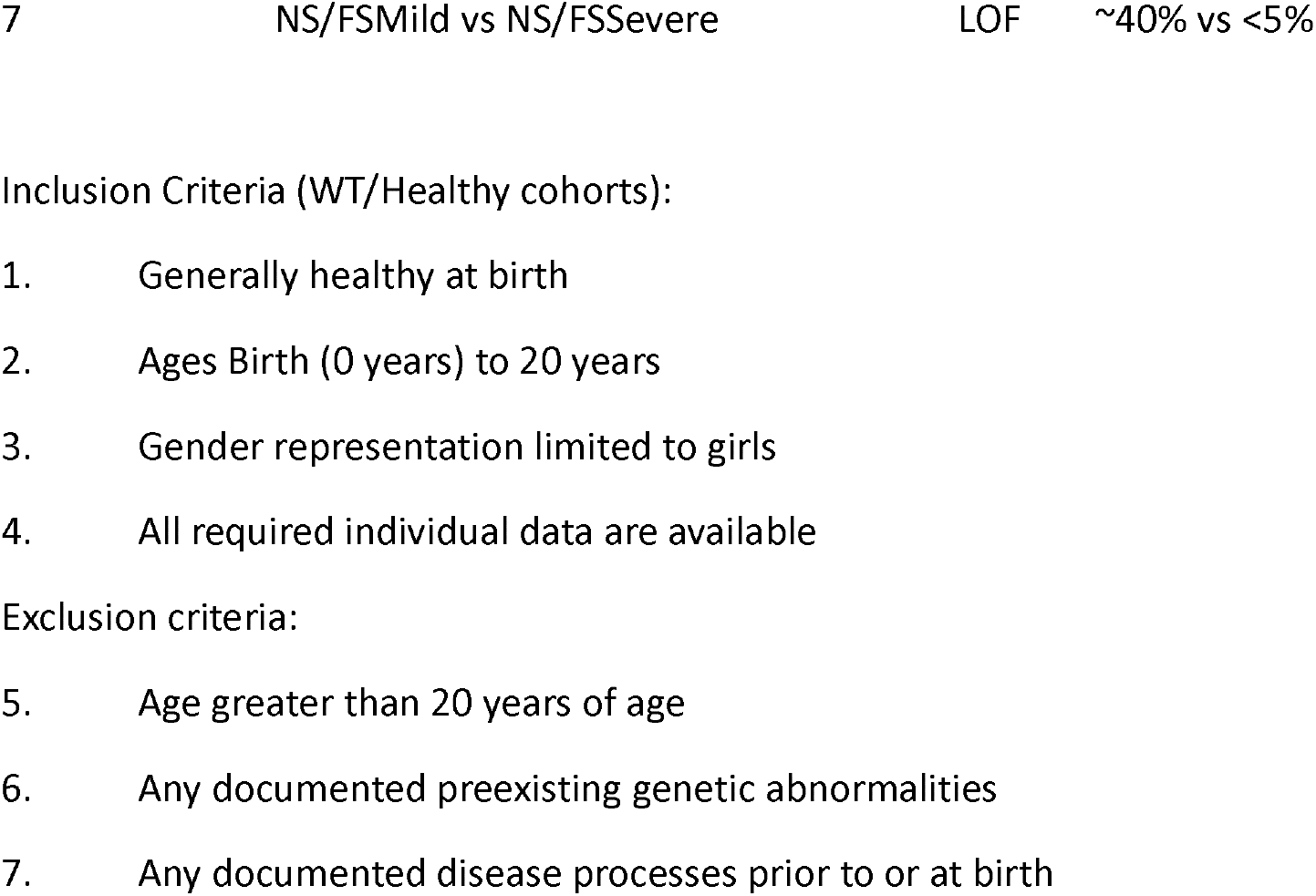
Summary of Cohorts, Comparisons, and Residual protein estimates for this virtual longitudinal study of HNRNPH2-NDD.

### The Affected Subjects

The HNRNPH2 mutations examined included: (i) the WT state, (ii) Gain of Function (GOF), (iii) Mild Missense (MS), (iv} Severe Missense (MS), (v) Mild Nonsense/Frameshift (NS/FS) and (vi) Severe Missense/Frameshift (NS/FS). The six age cohorts of twenty-five healthy subjects each underwent AI gene editing to introduce the Gain of Function (GOF) and loss of function (LOF) mutations for each of the six HNRNPH2 cohorts studied [13]. This process created thirty six highly matched cohorts where the only difference was the presence or absence of a specific gene mutation. In these well-matched cohorts, properties like obesity, hypertension and Type 2 Diabetes are emergent properties primarily associated with aging. The virtual approach has the major advantage that all subjects’ data were available for analysis since there was no attrition which would be common in traditional longitudinal studies of this kind. The data from all cohorts were evaluated beginning at birth (0 years) and continuing at 5-year intervals up to and including age 20 years of age (6 age cohorts).

The distinction between Mild and Severe HNRNPH2 mutations was based on estimates of residual normal functioning HNRNPH2 protein. For the Mild case, an estimate of ∼40% residual protein function was used and for the Severe case the estimated residual normal protein was <5%.

### Statistical Analysis

The Null hypothesis states that there are no statistically significant differences or at least medium effect sizes for the six affected groups compared to the age matched wild type (WT) subjects. The data was not normally distributed, so the non-parametric Wilcoxon signed rank test was used to estimate statistical significance. Given that multiple tests (N=17) were conducted, the conservative Bonferroni correction was applied. The corrected p value used to determine significance therefore became 0.05/17, or < 0.0029 for this study.

The alternative hypothesis states that there are significant differences in the affected groups compared to the healthy controls (WT).

The true effect size for the difference between affected groups and WT was estimated using Cliff’s delta (d). Cliff’s delta was used because the data were not normally distributed with a sample size of twenty-five subjects per cohort. To calculate Cliff’s delta the continuous data was transformed into interval data based on whether the data from the affected group was larger or smaller than the unaffected group. The Cliff’s delta was then calculated as (N (larger than) – N (smaller than))/the standard deviation of the differences between the groups [14,15]. This produced a range of effect size estimates between -1 (a large negative effect) and +1 (a large positive effect). A value close to zero was interpreted as having no effect. To determine the size of the effect we used the following heuristic scale: d <0.147 (negligible), d = 0.147 to <0.330 (small), d = 0.330 to <0.474 (medium) and d >= 0.474 (large) as suggested in [16]. To compensate for the modest sample size per cohort, all Cliff’s d values were modified using the Hedges correction [17] which was calculated to be 0.984. Final effect size estimates were obtained by multiplying the initial effect sizes by 0.984. The 95% CI around the effect size estimate was calculated using the Standard Error (SE) of the differences/square root of the sample size (2N data points).

## RESULTS

### (1) Wild Type (WT) subjects versus subjects with GOF mutations in HNRNPH2

The comparison between Wild Type (WT) subjects and those with Mild Missense (MSMild) mutations in HNRNPH2 revealed significant differences across multiple domains. The key findings are summarized below:

#### Neurological and Cognitive Functions

- Cognition/Memory/IQ: (all corrected p values are <0.0029) The estimated effect size of a GOF mutation in HNRNPH2 is maximally large and negative on Cognition/Memory/IQ (Cliff’s d = -0.984±0.000). This indicates that the GOF HNRNPH2 mutations are associated with impairment of Cognitive function that begins at birth and continues across all age cohorts studied.
- hCortMass: (all corrected p values are <0.0029 except age 20y) The estimated effect size of a GOF mutation in HNRNPH2 is maximally large and negative on brain Cortical Mass (Cliff’s d = -0.800±0.303). This indicates that the GOF HNRNPH2 mutations are associated with a degree of cortical atrophy or impaired development that begins at birth and continues across all age cohorts studied except for age 20 years.
- Speech & Language-Impaired: (all corrected p values are <0.0029) The estimated effect size of a GOF mutation in HNRNPH2 is maximally large and positive on Speech & Language impairment (Cliff’s d = 0.984±0.000). This indicates that the GOF HNRNPH2 mutations are associated with a degree of Speech & Language Impairment that begins at birth and persists across all age cohorts studied.
- Abnormal Behavior Issues: (all corrected p values are <0.0029) The estimated effect size of a GOF mutation in HNRNPH2 is maximally large and positive on Abnormal Behavioral issues (Cliff’s d = 0.984±0.000). This indicates that the GOF HNRNPH2 mutations are associated with a degree of Abnormal Behavioral issues that begin at birth and persist across all age cohorts studied.

#### Motor and Physical Functionality

- Motor functions/Normal: all corrected p values are <0.0029 Effects sizes are generally negative and large with an average d = -0.977±0.123 Normal motor functions are impaired in all age groups studied.
- Muscle Hypotonia: (all corrected p values are <0.0029 except for age 20 years) The estimated effect size of a GOF mutation in HNRNPH2 on Muscle Hypotonia is large and positive (Cliff’s d = 0.695±0.507). This indicates that the GOF HNRNPH2 mutations are associated with a degree of Muscle Hypotonia that begins at birth and persists across all age cohorts studied except for subjects aged 20 years.
- Ability to perform ADL: (all corrected p values are <0.0029) The estimated effect size of a GOF mutation in HNRNPH2 on the ability to perform ADL is consistently large and negative (Cliff’s d = -0.984±0.000). This indicates that the GOF HNRNPH2 mutations are associated with significant impairment of the ability to perform ADL that begins at birth and continues across all age cohorts studied.

#### Seizure and Sleep-Related Disorders

- Seizure Threshold: (all corrected p values are <0.0029) The estimated effect sizes of a GOF mutation in HNRNPH2 on Seizure threshold are consistently large and negative (Cliff’s d = -0.984±0.000). This indicates that the GOF HNRNPH2 mutations are associated with large decreases in the seizure threshold that begins at birth and continues across all age cohorts studied.
- Seizures Frequency: (all corrected p values are <0.0029) The estimated effect sizes of a GOF mutation in HNRNPH2 on Seizure Frequency are consistently large and positive (Cliff’s d = 0.984±0.000). This indicates that the GOF HNRNPH2 mutations are associated with significant increases in the seizure frequency that begins at birth and continues across all age cohorts studied.
- NeuroDevelopmental Disorder-Seizures: (all corrected p values are <0.0029) The estimated effect sizes of a GOF mutation in HNRNPH2 on Seizure Severity are consistently large and positive (Cliff’s d = 0.984±0.000). This indicates that the GOF HNRNPH2 mutations are associated with significant increases in the seizure severity that begins at birth and continues across all age cohorts studied.
- Sleep-Dysregulation/Loss: (all corrected p values are Significant at p<0.0029 except at Birth) The estimated effect sizes of a GOF mutation in HNRNPH2 on Sleep Dysregulation are variable and generally positive. Effect size is negligible (d = 0.000) at birth and then becomes moderate to large (d ≥ 0.472). The average effect size is 0.551±0.240.This indicates that the GOF HNRNPH2 mutations are associated with variable small to large sleep dysregulation beginning at age 2-3 years and continuing at all subsequent age groups studied.
- Gut-Brain Axis and Systemic Influence: GI-Gut-Brain Axis-Normal: (all corrected p values are <0.0029) Estimated effect sizes of a GOF mutation in HNRNPH2 on Normal Gut-Brain axis function are consistently large and negative (Cliff’s d = -0.984±0.000). This indicates that the GOF HNRNPH2 mutations are associated with large decreases in the Normal Gut-Brain axis function that begins at birth and continues across all age cohorts studied.

#### Genetic and Molecular Factors

- HNRNPH1: (all corrected p values are <0.0029 except at Birth and age 10 years) The estimated effect sizes of a GOF mutation in HNRNPH2 on HNRNPH1 protein are quite variable (average Cliff’s d = 0.039±0.642). GOF HNRNPH2 mutations appear to be associated with significant decreases in the HNRNPH1 protein that begins at age 2-3 years to age 10 years and then become large and negative (d < -0.826) in the oldest age groups. This interesting finding suggests an age related complex relationship HNRNPH2 GOF mutations and HNRNPH1 protein.
- HNRNPH2: (all corrected p values are <0.0029) The estimated effect sizes of a GOF mutation in HNRNPH2 on HNRNPH2 protein are consistently large and positive (Cliff’s d = 0.984±0.000) as expected. This indicates that the GOF HNRNPH2 mutations are associated with significant increases in HNRNPH2 protein that acquires GOF effects. This increase in HNRNPH2 protein begins at birth and continues across all age cohorts studied.

#### Quality of Life and Well-being

- QoL-Physical wellbeing (PW): (all corrected p values are <0.0029) The estimated effect sizes of a GOF mutation in HNRNPH2 on Physical Wellbeing are consistently large and negative (Cliff’s -d = -0.984±0.000). These results indicate that GOF HNRNPH2 mutations are associated with large negative effects on QofL-Physical Wellbeing that begins at birth and continues across all age cohorts studied.
- QoL-Psychological wellbeing (PsyW): (all corrected p values are <0.0029) The estimated effect sizes of a GOF mutation in HNRNPH2 on Psychological Wellbeing are consistently large and negative (Cliff’s -d = -0.984±0.000). These results indicate that GOF HNRNPH2 mutations are associated with large negative effects on QofL-Psychological Wellbeing that begins at birth and continues across all age cohorts studied.
- QoL-Social relationships (SR): (all corrected p values are <0.0029) The estimated effect sizes of a GOF mutation in HNRNPH2 on Social Relationships are consistently large and negative (Cliff’s -d = -0.984±0.000). These results indicate that GOF HNRNPH2 mutations are associated with large negative effects on QofL-Social Relationships that begin at birth and continue across all age cohorts studied.

These Results are summarized in Appendix B, Table 2

### (2) Wild Type (WT) subjects versus subjects with MSMild LOF mutations in HNRNPH2

The comparison between Wild Type (WT) subjects and those with Mild Missense (MSMild) mutations in HNRNPH2 revealed significant differences across multiple domains. The key findings are summarized below:

- Neurological and Cognitive Functions: Cognition/Memory/IQ: Subjects with MSMild mutations exhibited a significant decline in cognitive function compared to WT, with an average effect size of Cliff’s d = -0.984±0.000 (p < 0.0029). This indicates substantial cognitive deficits across all age cohorts. Cortical Mass (hCortMass): MSMild mutations were associated with a large reduction in cortical mass, with an effect size of Cliff’s d = -0.748±0.463 (p < 0.0029 except for age 20 years), suggesting potential cortical atrophy or impaired development. Speech & Language-Impaired: A large positive effect size (Cliff’s d = 0.945±0.053, p < 0.0029) was observed, indicating significant impairments in speech and language abilities across all cohorts. Abnormal Behavior Issues: MSMild mutations were linked to increased abnormal behavioral issues, with a large effect size of Cliff’s d = 0.984±0.000 (p < 0.0029).
- Motor and Physical Functionality: Motor functions: MSMild mutations resulted in a significant decline in motor functions, with an effect size of Cliff’s d = -0.873±0.070 (p < 0.0029). the decline in motor function is apparent in all age groups studied Muscle Hypotonia: A large positive effect size (Cliff’s d = 0.709±0.540, p < 0.0029 except at age 20 years) was observed, indicating consistent muscle weakness at all but the oldest age group studied. Ability to perform ADL: Significant impairment was observed in the ability to perform activities of daily living, with a large negative effect size of Cliff’s d = -0.984±0.000 (p < 0.0029) at all ages.
- Seizure and Sleep-Related Disorders: Seizure Threshold: A significant decrease in seizure threshold was noted (Cliff’s d = -0.984±0.000, p < 0.0029). Seizures Frequency: A significant increase in seizure frequency was observed (Cliff’s d = 0.984±0.000, p < 0.0029). NeuroDevelopmental Disorder-Seizures: The severity of seizures was significantly higher in the MSMild mutation group (Cliff’s d = 0.984±0.000, p < 0.0029). Sleep Dysregulation: Sleep dysregulation showed was variably impacted, with a small to medium effect size at age 10 year and beyond (Cliff’s d = 0.144±0.142, p > 0.0029 (NS, except age 10 years)).
- Gut-Brain Axis and Systemic Influence: GI-Gut-Brain Axis-Normal: A large negative effect size (Cliff’s d = -0.984±0.000, p < 0.0029) indicated widespread dysfunction in gut-brain communication pathways at all ages studied.
- Genetic and Molecular Factors: HNRNPH1: MSMild mutations led to a large decrease in HNRNPH1 protein levels across all age groups (average Cliff’s d = -0.984±0.000, p < 0.0029), suggesting potential downregulation of HNRNPH1 is associated with a MSMild mutation. HNRNPH2: A significant decrease in HNRNPH2 protein levels was observed consistent with the LOF mutation, with an effect size of Cliff’s d = -0.813±0.334 (p < 0.0029 except at age 20 years).
- Quality of Life and Well-being: Physical Wellbeing (PW): Significant decline in physical wellbeing in all age groups (Cliff’s d = -0.984±0.000, p < 0.0029). Psychological Wellbeing (PsyW): A large negative effect on psychological wellbeing was observed (Cliff’s d = -0.984±0.000, p < 0.0029). Social Relationships (SR): Social relationships were significantly impaired (Cliff’s d = - 0.984±0.000, p < 0.0029). These Results are summarized in Appendix B, Table 3

### (3) Wild Type (WT) subjects versus subjects with MSSevere LOF mutations in HNRNPH2

The comparison between Wild Type (WT) subjects and those with Severe Missense (MSSevere) mutations in HNRNPH2 revealed significant and consistent differences across multiple domains. The key findings are summarized below:

- Neurological and Cognitive Functions: Cognition/Memory/IQ: Subjects with MSSevere mutations exhibited a significant decline in cognitive function compared to WT, with an effect size of Cliff’s d = -0.984±0.000 (p < 0.0029). This indicates substantial cognitive deficits across all age cohorts. Cortical Mass (hCortMass): MSSevere mutations were associated with a large reduction in cortical mass, with an effect size of Cliff’s d = -0.984±0.000 (p < 0.0029), suggesting potential cortical atrophy or impaired development in all age groups. Speech & Language-Impaired: A large positive effect size (Cliff’s d = 0.984±0.000, p < 0.0029) was observed, indicating significant impairments in speech and language abilities across all cohorts. Abnormal Behavior Issues: MSSevere mutations were linked to increased abnormal behavioral issues, with an effect size of Cliff’s d = 0.984±0.000 (p < 0.0029) in all age groups.
- Motor and Physical Functionality: Motor functions: MSSevere mutations resulted in a significant decline in motor functions, with an effect size of Cliff’s d = -0.945±0.053 (p < 0.0029), across all age groups. Muscle Hypotonia: A large positive effect size (Cliff’s d = 0.984±0.000, p < 0.0029) was observed, indicating muscle weakness across all age groups. Ability to perform ADL: Significant impairment was observed in the ability to perform activities of daily living, with a large negative effect size of Cliff’s d = -0.984±0.000 (p < 0.0029).
- Seizure and Sleep-Related Disorders: Seizure Threshold: A significant decrease in seizure threshold was noted (Cliff’s d = -0.984±0.000, p < 0.0029). Seizures Frequency: A significant increase in seizure frequency was observed (Cliff’s d = 0.984±0.000, p < 0.0029). NeuroDevelopmental Disorder-Seizures: The severity of seizures was significantly higher in all MSSevere mutation age group (Cliff’s d = 0.984±0.000, p < 0.0029). Sleep Dysregulation: Sleep dysregulation showed a large negative impact, with a large and negative effect sizes (Cliff’s d = 0.984, p < 0.0029), in all age groups.
- Gut-Brain Axis and Systemic Influence: GI-Gut-Brain Axis-Normal: A large negative effect size (Cliff’s d = -0.984±0.000, p < 0.0029) indicated widespread dysfunction in the gut-brain communication pathway.
- Genetic and Molecular Factors: HNRNPH1: MSSevere mutations led to a significant decrease in HNRNPH1 protein levels (Cliff’s d = -0.984±0.000, p < 0.0029), suggesting potential downregulation in the context of a Severe mutation in HNRNPH2. HNRNPH2: A significant decrease in HNRNPH2 protein levels was observed across all age groups, with an effect size of Cliff’s d = -0.984±0.000 (p < 0.0029).
- Quality of Life and Well-being: Physical Wellbeing (PW): Significant decline in physical wellbeing (Cliff’s d = -0.984±0.000, p < 0.0029). Psychological Wellbeing (PsyW): A large negative effect on psychological wellbeing was observed (Cliff’s d = -0.984±0.000, p < 0.0029). Social Relationships (SR): Social relationships were significantly impaired (Cliff’s d = - 0.984±0.000, p < 0.0029). These Results are summarized in Appendix B, Table 4

### (4) Wild Type (WT) subjects versus subjects with NS/FSMild LOF mutations in HNRNPH2

The comparison between Wild Type (WT) subjects and those with Nonsense/Frameshift Mild (NS/FSMild) mutations in HNRNPH2 revealed significant differences across multiple domains. The key findings are summarized below:

- Neurological and Cognitive Functions: Cognition/Memory/IQ: Subjects with NS/FSMild mutations exhibited a significant decline in cognitive function compared to WT, with an effect size of Cliff’s d = -0.984±0.000 (p < 0.0029). This indicates substantial cognitive deficits across all age cohorts. Cortical Mass (hCortMass): NS/FSMild mutations were associated with a large reduction in cortical mass, with an effect size of Cliff’s d = -0.748±0.316 (p < 0.0029 except age 20 years), suggesting potential cortical atrophy or impaired development that begins at Birth. Speech & Language-Impaired: A large positive effect size (Cliff’s d = 0.945±0.053, p < 0.0029) was observed, indicating significant impairments in speech and language abilities across all cohorts. Abnormal Behavior Issues: NS/FSMild mutations were linked to increased abnormal behavioral issues, with an effect size of Cliff’s d = 0.984±0.000 (p < 0.0029).
- Motor and Physical Functionality: Motor functions: NS/FSMild mutations resulted in a significant decline in motor functions, with an average effect size of Cliff’s d = -0.905±0.069 (p < 0.0029), showing very slight variability with age. Muscle Hypotonia: A large positive effect size (Cliff’s d = 0.984±0.000, p < 0.0029 except at age 20 years) was observed, from Birth to age 15 years indicating consistent muscle weakness. Ability to perform ADL: Significant impairment was observed in the ability to perform activities of daily living, with a large negative effect size of Cliff’s d = -0.984±0.000 (p < 0.0029) in all age groups.
- Seizure and Sleep-Related Disorders: Seizure Threshold: A significant decrease in seizure threshold was noted (Cliff’s d = -0.984±0.000, p < 0.0029). Seizures Frequency: A significant increase in seizure frequency was observed (Cliff’s d = 0.984±0.000, p < 0.0029). NeuroDevelopmental Disorder-Seizures: The severity of seizures was significantly higher in the NS/FSMild mutation groups (Cliff’s d = 0.984±0.000, p < 0.0029). Sleep Dysregulation: Sleep dysregulation showed a varied negligible impact, with a medium positive effect size at age 10 years (Cliff’s d = 0.072, p > 0.0029 except at 10 years of age).
- Gut-Brain Axis and Systemic Influence: GI-Gut-Brain Axis-Normal: A large negative effect size (Cliff’s d = -0.984±0.000, p < 0.0029) indicated widespread dysfunction in the gut-brain communication pathway.
- Genetic and Molecular Factors: HNRNPH1: NS/FSMild mutations led to a significant decrease in HNRNPH1 protein levels (Cliff’s d = -0.984±0.000, p < 0.0029), suggesting potential downregulation. HNRNPH2: A significant decrease in HNRNPH2 protein levels was observed, with an effect size of Cliff’s d = -0.984±0.000 (p < 0.0029).
- Quality of Life and Well-being: Physical Wellbeing (PW): Significant decline in physical wellbeing was observed (Cliff’s d = - 0.984±0.000, p < 0.0029). Psychological Wellbeing (PsyW): A large negative effect on psychological wellbeing was observed (Cliff’s d = -0.984±0.000, p < 0.0029). Social Relationships (SR): Social relationships were significantly impaired (Cliff’s d = -0.984, p < 0.0029). These Results are summarized in Appendix B, Table 5

### (5) Wild Type (WT) subjects versus subjects with NS/FSSevere LOF mutations in HNRNPH2

The comparison between Wild Type (WT) subjects and those with Nonsense/Frameshift Severe (NS/FSSevere) mutations in HNRNPH2 revealed significant differences across multiple domains. The key findings are summarized below:

- Neurological and Cognitive Functions: Cognition/Memory/IQ: Subjects with NS/FSSevere mutations exhibited a significant decline in cognitive function compared to WT, with an effect size of Cliff’s d = -0.984±0.000 (p < 0.0029). This indicates substantial cognitive deficits across all age cohorts. Cortical Mass (hCortMass): NS/FSSevere mutations were associated with a large reduction in cortical mass, with an effect size of Cliff’s d = -0.984±0.000 (p < 0.0029), suggesting potential cortical atrophy or impaired development. Speech & Language-Impaired: A large positive effect size (Cliff’s d = 0.984±0.000, p < 0.0029) was observed, indicating significant impairments in speech and language abilities across all cohorts. Abnormal Behavior Issues: NS/FSSevere mutations were linked to increased abnormal behavioral issues, with an effect size of Cliff’s d = 0.984±0.000 (p < 0.0029).
- Motor and Physical Functionality: Motor functions: NS/FSSevere mutations resulted in a significant decline in motor functions, with an effect size of Cliff’s d = -0.945±0.053 (p < 0.11), across all ages studied. Muscle Hypotonia: A large positive effect size (Cliff’s d = 0.984±0.000, p < 0.0029) was observed, indicating consistent muscle weakness. Ability to perform ADL: Significant impairment was observed in the ability to perform activities of daily living, with a large negative effect size of Cliff’s d = -0.984±0.000 (p < 0.0029).
- Seizure and Sleep-Related Disorders: Seizure Threshold: A significant decrease in seizure threshold was noted (Cliff’s d = -0.984±0.000, p < 0.0029). Seizures Frequency: A significant increase in seizure frequency was observed (Cliff’s d = 0.984±0.000, p < 0.0029). NeuroDevelopmental Disorder-Seizures: The severity of seizures was significantly higher in the NS/FSSevere mutation group (Cliff’s d = 0.984±0.000, p < 0.0029). Sleep Dysregulation: Sleep dysregulation showed an average impact, with an average large effect (Cliff’s d = 0.925±0.039, p < 0.0029).
- Gut-Brain Axis and Systemic Influence: GI-Gut-Brain Axis-Normal: A large negative effect size (Cliff’s d = -0.984±0.000, p < 0.0029) indicated widespread dysfunction in the gut-brain communication pathway.
- Genetic and Molecular Factors: HNRNPH1: NS/FSSevere mutations led to a significant decrease in HNRNPH1 protein levels (Cliff’s d = -0.984±0.000, p < 0.0029), suggesting potential downregulation. HNRNPH2: A significant decrease in HNRNPH2 protein levels was observed, with an effect size of Cliff’s d = -0.984±0.000 (p < 0.0029).
- Quality of Life and Well-being: Physical Wellbeing (PW): Significant decline in physical wellbeing was observed (Cliff’s d = - 0.984±0.000, p < 0.0029). Psychological Wellbeing (PsyW): A large negative effect on psychological wellbeing was observed (Cliff’s d = -0.984±0.000, p < 0.0029). Social Relationships (SR): Social relationships were significantly impaired (Cliff’s d = - 0.984±0.000, p < 0.0029). These Results are summarized in Appendix B, Table 6

### (6) Comparing Subjects with MSSevere versus MSMild Mutations

The comparison between Severe Missense (MSSevere) and Mild Missense (MSMild) mutations in HNRNPH2 serves as a quality control to ensure consistent and significant differentiation between the severity of mutations. The key findings are summarized below:

- Neurological and Cognitive Functions: Cognition/Memory/IQ: The MSSevere group showed a significant decline in cognitive function compared to the MSMild group, with an effect size of Cliff’s d = -0.984 (p < 0.0029). This indicates that the cognitive deficits are markedly worse in the severe mutation group. Cortical Mass (hCortMass): There was a significant reduction in cortical mass in the MSSevere group compared to MSMild, beginning at age 2-3 years with an average effect size of Cliff’s d = - 0.643±0.459 (all p values < 0.0029). A large positive effect size is observed at birth (d = 0.512±0.459) before becoming consistently negative. Speech & Language-Impaired: A positive effect size (Cliff’s d = 0.984±0.000, p < 0.0029) indicates significantly worse impairments in speech and language abilities in the MSSevere group. Abnormal Behavior Issues: The MSSevere group was linked to a significant increase in abnormal behavioral issues compared to the MSMild group, with an effect size of Cliff’s d = 0.984±0.000 (p < 0.0029).
- Motor and Physical Functionality: Motor functions: The MSSevere group exhibited a more significant decline in motor functions compared to the MSMild group, with an effect size of Cliff’s d = -0.866±0.173 (p < 0.0029 except at age 20 years). Muscle Hypotonia: The effect size is variably negative prior to age 10 years (d = -0.197 to -0.827) A large and positive effect size (Cliff’s d = 0.984, p < 0.0029) was observed from age 10 to 20 years, indicating that muscle weakness becomes significantly more severe in older MSSevere groups. Corrected p values were <0.0029 except at Birth and age 5 years. Ability to perform ADL: The ability to perform activities of daily living was significantly worse in the MSSevere groups, with a large negative effect size of Cliff’s d = -0.984±0.000 (p < 0.0029).
- Seizure and Sleep-Related Disorders: Seizure Threshold: A significant decrease in seizure threshold was noted in the MSSevere group compared to MSMild, with Cliff’s d = -0.984±0.000 (p < 0.0029). Seizures Frequency: The MSSevere group experienced a significant increase in seizure frequency compared to MSMild, with an effect size of Cliff’s d = 0.984±0.000 (p < 0.0029). NeuroDevelopmental Disorder-Seizures: The severity of seizures was significantly higher in the MSSevere group, with an effect size of Cliff’s d = 0.984 (p < 0.0029). Sleep Dysregulation: Sleep dysregulation showed a large effect size, in the MSSevere groups (Cliff’s d = 0.984±0.000, p < 0.0029).
- Gut-Brain Axis and Systemic Influence: GI-Gut-Brain Axis-Normal: A large negative effect size (Cliff’s d = -0.984±0.000, p < 0.0029) indicated more significant dysfunction in the gut-brain communication pathway in the MSSevere group.
- Genetic and Molecular Factors: HNRNPH1: The MSSevere group had a significantly greater reduction in HNRNPH1 protein levels compared to the MSMild group (Cliff’s d = -0.984±0.000, p < 0.0029). HNRNPH2: A significant decrease in HNRNPH2 protein levels was observed in the MSSevere group, with an effect size of Cliff’s d = -0.984±0.000 (p < 0.0029).
- Quality of Life and Well-being: Physical Wellbeing (PW): Physical wellbeing was significantly worse in the MSSevere group compared to MSMild, with a large negative effect size of Cliff’s d = -0.984±0.000 (p < 0.0029). Psychological Wellbeing (PsyW): A large negative effect on psychological wellbeing was observed in the MSSevere group, with an effect size of Cliff’s d = -0.984±0.000 (p < 0.0029). Social Relationships (SR): Social relationships were significantly impaired in the MSSevere group, with an effect size of Cliff’s d = -0.984±0.000 (p < 0.0029). These Results are summarized in Appendix B, Table 7

### (7) Comparing Subjects with NS/FSSevere versus NS/FSMild Mutations

The comparison between Severe Nonsense/Frameshift (NS/FSSevere) and Mild Nonsense/Frameshift (NS/FSMild) mutations in HNRNPH2 revealed significant differences, emphasizing the distinction between the severity of these mutations. The key findings are summarized below:

- Neurological and Cognitive Functions: Cognition/Memory/IQ: The NS/FSSevere group exhibited a significant decline in cognitive function compared to the NS/FSMild group, with an effect size of Cliff’s d = -0.984±0.000 (p < 0.0029). This indicates that cognitive deficits are more pronounced in the severe mutation group. Cortical Mass (hCortMass): A significant reduction in cortical mass was observed in the NS/FSSevere group compared to NS/FSMild, with a large effect size of Cliff’s d = -0.853±0.103 (p < 0.0029). Speech & Language-Impaired: The NS/FSSevere group showed significantly worse impairments in speech and language abilities, with a positive effect size of Cliff’s d = 0.984±0.000 (p < 0.0029). Abnormal Behavior Issues: Increased abnormal behavioral issues were noted in the NS/FSSevere group compared to NS/FSMild, with an average large effect size of Cliff’s d = 0.945±0.049 (p < 0.0029).
- Motor and Physical Functionality: Motor functions: The NS/FSSevere group exhibited a more significant decline in motor functions compared to the NS/FSMild group, with an effect size of Cliff’s d = -0.984±0.000 (p < 0.0029). Muscle Hypotonia: Muscle weakness was significantly more severe in the NS/FSSevere group, with a positive effect size of Cliff’s d = 0.840±0.253 (p < 0.0029 except at age 5 years). Ability to perform ADL: The ability to perform activities of daily living was significantly worse in the NS/FSSevere group, with a large negative effect size of Cliff’s d = -0.984±0.000 (p < 0.0029).
- Seizure and Sleep-Related Disorders: Seizure Threshold: A significant decrease in seizure threshold was noted in the NS/FSSevere group compared to NS/FSMild, with Cliff’s d = -0.984±0.000 (p < 0.0029). Seizures Frequency: The NS/FSSevere group experienced a significant increase in seizure frequency compared to NS/FSMild, with an effect size of Cliff’s d = 0.984±0.000 (p < 0.0029). NeuroDevelopmental Disorder-Seizures: The severity of seizures was significantly higher in the NS/FSSevere group, with an effect size of Cliff’s d = 0.984±0.000 (p < 0.0029). Sleep Dysregulation: Sleep dysregulation showed a large positive impact, with a medium but still significant effect size observed at 10 years in the NS/FSSevere group vs the NS/FS mild group (average Cliff’s d = 0.859±0.155, p < 0.0029).
- Gut-Brain Axis and Systemic Influence: GI-Gut-Brain Axis-Normal: A large negative effect size (Cliff’s d = -0.984±0.000, p < 0.0029) indicated more significant dysfunction in the gut-brain communication pathway in the NS/FSSevere group.
- Genetic and Molecular Factors: HNRNPH1: The NS/FSSevere group had a significantly greater reduction in HNRNPH1 protein levels compared to the NS/FSMild group (Cliff’s d = -0.984±0.000, p < 0.0029). HNRNPH2: A significant decrease in HNRNPH2 protein levels was observed in the NS/FSSevere group, with an effect size of Cliff’s d = -0.984±0.000 (p < 0.0029).
- Quality of Life and Well-being: Physical Wellbeing (PW): Physical wellbeing was significantly worse in the NS/FSSevere group compared to NS/FSMild, with a maximally large negative effect size of Cliff’s d = -0.984±0.000 (p < 0.0029). Psychological Wellbeing (PsyW): A large negative effect on psychological wellbeing was observed in the NS/FSSevere group, with an effect size of Cliff’s d = -0.984±0.000 (p < 0.0029). Social Relationships (SR): Social relationships were significantly impaired in the NS/FSSevere group, with an effect size of Cliff’s d = -0.984±0.000 (p < 0.0029). These Results are summarized in Appendix B, Table 8

## Discussion

This study provides significant insights into the functional consequences of Mild and Severe HNRNPH2 mutations, particularly distinguishing between gain-of-function (GOF) and loss-of-function (LOF) effects. By utilizing advanced aiHumanoid simulations, we conducted a detailed virtual longitudinal analysis that replicates the systemic and neurological impacts of HNRNPH2 mutations from birth through young adulthood.

### Key Findings and Implications

#### 1. Gain of Function Mutations if HNRNPH2

The analysis of Gain-of-Function (GOF) mutations in HNRNPH2 reveals a distinctive and severe effect on neurodevelopmental outcomes, emphasizing the critical role of these mutations in the pathophysiology of HNRNPH2-related NDDs. Our results indicate that GOF mutations are associated with profound cognitive deficits, significant reductions in cortical mass, and severe impairments in motor function, consistent with findings from similar studies on other neurodevelopmental disorders, such as those described by Kelvington and Abel [2], where RNA granule dysregulation was identified as a key mechanism. Furthermore, the increased seizure frequency and lowered seizure threshold observed in GOF cohorts mirror the neurological disorders seen in ARX-related epilepsy and Rett syndrome. These findings suggest that there are shared neural hyperexcitability pathways [1]. The variable changes in HNRNPH1 protein levels observed in GOF mutations further complicate the clinical picture, suggesting potential age-dependent compensatory mechanisms or feedback loops, which may exacerbate the phenotypic severity as described by Korff et al. [6] in mouse models. These findings emphasize the importance of developing targeted therapeutic strategies that specifically address the overactive pathways induced by GOF mutations, potentially through RNA-based interventions or small molecules designed to modulate HNRNP protein function [7].

#### 2. Dominant-Negative Effects of LOF Mutations

The findings from LOF mutations, including both missense and nonsense/frameshift mutations, strongly suggest a dominant-negative effect. The severe impairment observed in cognitive function, motor abilities, and other systemic markers in the LOF cohorts—especially the MSSevere and NS/FSSevere groups—aligns with recent research highlighting the potential for mutant HNRNPH2 proteins to disrupt not only their function but also the compensatory mechanisms provided by HNRNPH1 [18, 19]. This disruption supports the hypothesis that these mutations may exert a toxic gain-of-function effect or interfere with the genetic compensation by HNRNPH1, leading to the profound phenotypic consequences observed [20,21].

Mechanistically, the dominant-negative effects likely involve the interference of mutant HNRNPH2 protein with the normal function of wild-type HNRNPH2 or other hnRNP proteins, such as HNRNPH1. This interference could disrupt critical RNA processing functions, leading to widespread negative effects on gene expression and cellular function [22]. The reduced ability of HNRNPH1 to compensate in the presence of mutant HNRNPH2, as suggested by recent murine models, underscores the importance of exploring these protein-protein interactions in greater detail (19, 21).

Importantly, this study helps to extend our existing knowledge by providing the best evidence to date of dominant-negative effects, which had been hypothesized but not rigorously demonstrated in the context of HNRNPH2-related NDD [18,20]. This insight is critical for understanding the pathophysiology of the disorder and for guiding the development of targeted therapies [24, 26].

#### 3. Comparative Severity of Mutations

Our data clearly differentiate the impact of mild versus severe mutations. In each category (MS and NS/FS), the severe mutations resulted in significantly worse outcomes across almost all measured domains, including cognitive, motor, behavioral, and quality of life indicators [20,23]. These findings underscore the importance of understanding the gradations in mutation severity because they directly impact prognosis and therapeutic targeting [21].

Understanding the differential impacts of mutation severity is critical for developing personalized treatment strategies. Children with severe mutations may benefit from more aggressive therapeutic interventions early in development, while children with mild mutations might be managed with less intensive approaches [24]. These distinctions also underscore the need for detailed genetic analysis to clarify mutational status in clinical settings to inform treatment decisions [25].

#### 4. Seizure Findings and Implications

Our aiHumanoid simulations revealed a significant association between HNRNPH2 mutations and increased seizure activity, with particular emphasis on seizure threshold and frequency. Specific mutations, particularly the more severe LOF variants, were linked to a lower seizure threshold and higher seizure frequency, consistent with findings in similar neurodevelopmental disorders like ARX-related epilepsy [30] and Rett syndrome [29]. These results suggest that HNRNPH2 mutations disrupt neural circuits, reducing the seizure threshold and the brain’s ability to prevent seizure onset.

The observed variation in seizure frequency across different mutation types underscores the importance of early and precise assessment in affected individuals. Monitoring seizure threshold and frequency could guide personalized therapeutic strategies, aiming to stabilize neural activity and reduce seizure occurrence [47]. This is particularly relevant given the shared molecular pathways between epilepsy and NDDs, such as those highlighted in recent studies on autism spectrum disorder [51]. This approach aligns with the understanding that seizure frequency is a critical prognostic indicator in epilepsy [39]. Future research should explore targeted interventions that address the specific pathways involved in seizure generation in HNRNPH2-related NDD, potentially improving outcomes for patients [24].

#### 5. Potential for Therapeutic Interventions

The consistent patterns of impairment observed across different cohorts suggest potential therapeutic windows. The early onset of cognitive deficits and motor impairments, along with the progressive deterioration seen in severe mutations, points to the necessity of early intervention strategies [22]. Furthermore, the upregulation of HNRNPH1 in knockout mouse models raises the possibility that enhancing this compensatory mechanism might offer a viable therapeutic avenue [21,22].

Future research should focus on strategies to enhance HNRNPH1 expression or function in patients with HNRNPH2 LOF mutations. Small molecules or gene therapies that can boost HNRNPH1 levels or stabilize its function could mitigate the dominant-negative effects of mutant HNRNPH2 [25,26]. Additionally, exploring the potential for RNA-based therapies to selectively inhibit the expression of mutant HNRNPH2 could provide another avenue for treatment [22].

#### 6. Integration of AI Assisted Virtual Simulations in NDD Research

This study illustrates the potential of aiHumanoid simulations for modeling complex neurodevelopmental disorders. By integrating genetic, cellular, and molecular data, we can predict the longitudinal outcomes of various mutations, providing a robust platform for future virtual drug trials [23]. This approach not only accelerates the pace of discovery but also reduces the ethical and logistical challenges associated with traditional longitudinal studies [20].

While the aiHumanoid simulations offer several significant advantages, it is important to validate these findings in biological models. Future studies should aim to corroborate the predictions made by these AI-informed simulations using in vitro and in vivo systems [21]. This will ensure that the insights gained from virtual models are applicable to real-world clinical scenarios [25].

#### 7. Comparison to Our Current Understanding of HNRNPH2-NDD

The findings from this virtual study add several new dimensions to the current understanding of HNRNPH2-related neurodevelopmental disorders (NDDs). Historically, HNRNPH2 mutations have been associated with a spectrum of developmental delays, motor deficits, and cognitive impairments, as highlighted in studies by Gillentine et al. and Kelvington et al. [18,19]. However, much of the early research focused on characterizing the phenotypic spectrum and basic molecular mechanisms, without delving deeply into the comparative impacts of different types of mutations (e.g., GOF vs. LOF, or mild vs. severe mutations) [24,25].

#### 8. Ethical Considerations Regarding the Use of aiHumanoid Simulations

The use of aiHumanoid simulations to study HNRNPH2-related NDDs, particularly in multiple all-female virtual cohorts, raises specific ethical considerations. The primary concern is ensuring that these simulations accurately reflect the biological and clinical realities of HNRNPH2 mutations. While simulations are a powerful tool for exploring long-term outcomes and potential therapies, their predictions must be rigorously validated, potentially through comparison with actual clinical data, to avoid any misguidance in future research or clinical applications. The use of all-female cohorts, necessitated by the X-linked nature of HNRNPH2 mutations, emphasizes the importance of tailoring simulations to specific genetic contexts, ensuring that findings are relevant and applicable. Moreover, the use of AI-generated population-based subjects, without reliance on actual patient data, significantly minimizes ethical concerns related to privacy and consent.

In conclusion, the findings from this virtual study contribute to a deeper understanding of the pathophysiology of HNRNPH2-related neurodevelopmental disorders. They emphasize the need for early diagnosis and personalized interventions, particularly for severe LOF mutations that may produce dominant-negative effects [23, 25]. Future research should focus on therapeutic strategies that can either mitigate these dominant-negative effects or enhance compensatory mechanisms, such as the upregulation of HNRNPH1 [22,21]. Finally, these results lay a solid foundation for supporting early stage disease modifying aiHumanoid virtual drugs trials like [12].

## Data Availability

All data produced in the present study are available upon reasonable request to the authors

### Appendix A: HNRNPH2-NDD Feature References

1. Able to Perform ADL (Activities of Daily Living) [2,27]
2. Abnormal Behavior Issues [28,29]
3. Cognition/Memory/IQ [7,30]
4. GI-Gut-Brain Axis Normal [31,32]
5. Cortical Mass [33.34]
6. HNRNPH1 [7,35]
7. HNRNPH2 [2,36]
8. Motor Functions/Normal [29,37]
9. Muscle Hypotonia [7,38]
10. Neurodevelopmental Disorder Seizures [2,39,51]
11. QoL-Physical Wellbeing (PW) [40,41]
12. QoL-Psychological Wellbeing (PsyW) [42,43]
13. QoL-Social Relationships (SR) [44,45]
14. Seizure Threshold [29,46]
15. Seizure Frequency [7,47]
16. Sleep-Dysregulation/Loss [48,49]
17. Speech & Language-Impaired [29,50]

### Appendix B: : Cliff’s delta (d) Effect Sizes and Significance Levels for Comparison of WT vs Various HNRNPH2 mutations

**Table 2:** Effect Sizes and Significance Levels for Comparison of WT vs GOF HNRNPH2 mutations.

|  |  |  |  |
| --- | --- | --- | --- |
| Outcome | Effect Size (Cliff's d) | Significance (corrected p-value) | Direction of Effect |
| Ability to perform ADL | -0.984 ± 0.000 | <0.0029 | Negative |
| Abnormal Behavior Issues | 0.984 ± 0.000 | <0.0029 | Positive |
| Cognition/Memory/IQ | -0.984 ± 0.000 | <0.0029 | Negative |
| GI-Gut-Brain Axis-Normal | -0.984 ± 0.000 | <0.0029 | Negative |
| hCortMass | -0.800 ± 0.303 | <0.0029 (except 20y) | Negative |
| HNRNPH1 | 0.039 ± 0.642 | <0.0029 (except Birth&10y) | Neutral |
| HNRNPH2 | 0.984 ± 0.000 | <0.0029 | Positive |
| Motor functions/Normal | -0.977 ± 0.113 | <0.0029 | Negative |
| Muscle Hypotonia | 0.695 ± 0.507 | <0.0029 (except 20y) | Positive |
| NeuroDevelopmental Disorder-Seizures | 0.984 ± 0.000 | <0.0029 | Positive |
| QoL-Physical wellbeing (PW) | -0.984 ± 0.000 | <0.0029 | Negative |
| QoL-Psychological wellbeing (PsyW) | -0.984 ± 0.000 | <0.0029 | Negative |
| QoL-Social relationships (SR) | -0.984 ± 0.000 | <0.0029 | Negative |
| Seizure Threshold | -0.984 ± 0.000 | <0.0029 | Negative |
| Seizures Frequency | 0.984 ± 0.000 | <0.0029 | Positive |
| Sleep-Dysregulation/Loss | 0.551 ± 0.240 | <0.0029 (except at Birth) | Positive (variable) |
| Speech & Language-Impaired | 0.984 ± 0.000 | <0.0029 | Positive |

**Table 3:**
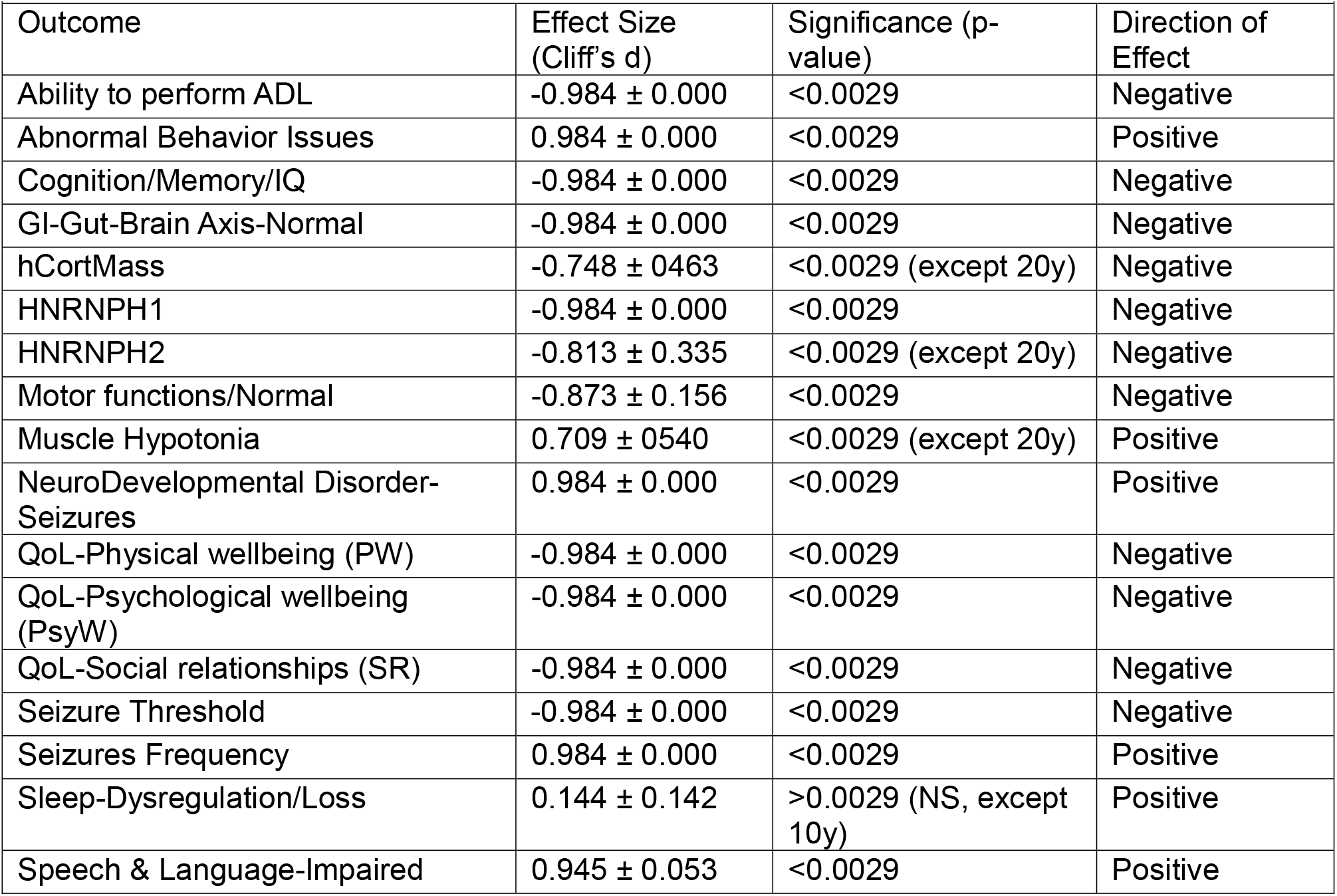
Effect Sizes and Significance Levels for Comparison of WT vs. MSMild HNRNPH2 LOF mutations.

**Table 4:** Effect Sizes and Significance Levels for Comparison of WT vs. MSSevere HNRNPH2 LOF mutations.

|  |  |  |  |
| --- | --- | --- | --- |
| Outcome | Effect Size (Cliff's d) | Significance (p-value) | Direction of Effect |
| Ability to perform ADL | -0.984 ± 0.000 | <0.0029 | Negative |
| Abnormal Behavior Issues | 0.984 ± 0.000 | <0.0029 | Positive |
| Cognition/Memory/IQ | -0.984 ± 0.000 | <0.0029 | Negative |
| GI-Gut-Brain Axis-Normal | -0.984 ± 0.000 | <0.0029 | Negative |
| hCortMass | -0.984 ± 0.000 | <0.0029 | Negative |
| HNRNPH1 | -0.984 ± 0.000 | <0.0029 | Negative |
| HNRNPH2 | -0.984 ± 0.000 | <0.0029 | Negative |
| Motor functions/Normal | -0.945 ± 0.053 | <0.0029 | Negative |
| Muscle Hypotonia | 0.984 ± 0.000 | <0.0029 | Positive |
| NeuroDevelopmental Disorder-Seizures | 0.984 ± 0.000 | <0.0029 | Positive |
| QoL-Physical wellbeing (PW) | -0.984 ± 0.000 | <0.0029 | Negative |
| QoL-Psychological wellbeing (PsyW) | -0.984 ± 0.000 | <0.0029 | Negative |
| QoL-Social relationships (SR) | -0.984 ± 0.000 | <0.0029 | Negative |
| Seizure Threshold | -0.984 ± 0.000 | <0.0029 | Negative |
| Seizures Frequency | 0.984 ± 0.000 | <0.0029 | Positive |
| Sleep-Dysregulation/Loss | 0.984 ± 0.000 | <0.0029 | Positive |
| Speech & Language-Impaired | 0.984 ± 0.000 | <0.0029 | Positive |

**Table 5:** Effect Sizes and Significance Levels for Comparison of WT vs. NS/FSMild HNRNPH2 mutations.

|  |  |  |  |
| --- | --- | --- | --- |
| Outcome | Effect Size (Cliff's d) | Significance (p-value) | Direction of Effect |
| Ability to perform ADL | -0.984 ± 0.000 | <0.0029 | Negative |
| Abnormal Behavior Issues | -0.984 ± 0.000 | <0.0029 | Positive |
| Cognition/Memory/IQ | -0.984 ± 0.000 | <0.0029 | Negative |
| GI-Gut-Brain Axis-Normal | -0.984 ± 0.000 | <0.0029 | Negative |
| hCortMass | -0.748 ± 0.316 | <0.0029 (except 20y) | Negative |
| HNRNPH1 | -0.984 ± 0.000 | <0.0029 | Negative |
| HNRNPH2 | -0.984 ± 0.000 | <0.0029 | Negative |
| Motor functions/Normal | -0.905 ± 0.069 | <0.0029 | Negative |
| Muscle Hypotonia | 0.722 ± 0.514 | <0.0029 | Positive |
| NeuroDevelopmental Disorder-Seizures | -0.984 ± 0.000 | <0.0029 | Positive |
| QoL-Physical wellbeing (PW) | -0.984 ± 0.000 | <0.0029 | Negative |
| QoL-Psychological wellbeing (PsyW) | -0.984 ± 0.000 | <0.0029 | Negative |
| QoL-Social relationships (SR) | -0.984 ± 0.000 | <0.0029 | Negative |
| Seizure Threshold | -0.984 ± 0.000 | <0.0029 | Negative |
| Seizures Frequency | -0.984 ± 0.000 | <0.0029 | Positive |
| Sleep-Dysregulation/Loss | 0.072 ± 0.141 | >0.0029 (except 10y) | Neutral |
| Speech & Language-Impaired | -0.984 ± 0.000 | <0.0029 | Positive |

**Table 6:** Effect Sizes and Significance Levels for Comparison of WT vs. NS/FSSevere HNRNPH2 LOF mutations.

|  |  |  |  |
| --- | --- | --- | --- |
| Outcome | Effect Size (Cliff's d) | Significance (p-value) | Direction of Effect |
| Ability to perform ADL | -0.984 ± 0.000 | <0.0029 | Negative |
| Abnormal Behavior Issues | -0.984 ± 0.000 | <0.0029 | Positive |
| Cognition/Memory/IQ | -0.984 ± 0.000 | <0.0029 | Negative |
| GI-Gut-Brain Axis-Normal | -0.984 ± 0.000 | <0.0029 | Negative |
| hCortMass | -0.984 ± 0.000 | <0.0029 | Negative |
| HNRNPH1 | -0.984 ± 0.000 | <0.0029 | Negative |
| HNRNPH2 | -0.984 ± 0.000 | <0.0029 | Negative |
| Motor functions/Normal | -0.945 ± 0.053 | <0.0029 | Negative |
| Muscle Hypotonia | -0.984 ± 0.000 | <0.0029 | Positive |
| NeuroDevelopmental Disorder-Seizures | -0.984 ± 0.000 | <0.0029 | Positive |
| QoL-Physical wellbeing (PW) | -0.984 ± 0.000 | <0.0029 | Negative |
| QoL-Psychological wellbeing (PsyW) | -0.984 ± 0.000 | <0.0029 | Negative |
| QoL-Social relationships (SR) | -0.984 ± 0.000 | <0.0029 | Negative |
| Seizure Threshold | -0.984 ± 0.000 | <0.0029 | Negative |
| Seizures Frequency | -0.984 ± 0.000 | <0.0029 | Positive |
| Sleep-Dysregulation/Loss | 0.925 ± 0.389 | <0.0029 | Positive |
| Speech & Language-Impaired | -0.984 ± 0.000 | <0.0029 | Positive |

**Table 7:** Effect Sizes and Significance Levels for comparison of MSSevere vs. MSMild HNRNPH2 mutations.

|  |  |  |  |
| --- | --- | --- | --- |
| Outcome | Effect Size (Cliff's d) | Significance (p-value) | Direction of Effect |
| Ability to perform ADL | -0.984 ± 0.000 | <0.0029 | Negative |
| Abnormal Behavior Issues | 0.984 ± 0.000 | <0.0029 | Positive |
| Cognition/Memory/IQ | -0.984 ± 0.000 | <0.0029 | Negative |
| GI-Gut-Brain Axis-Normal | -0.984 ± 0.000 | <0.0029 | Negative |
| hCortMass | -0.643 ± 0.459 | <0.0029 | Negative |
| HNRNPH1 | -0.984 ± 0.000 | <0.0029 | Negative |
| HNRNPH2 | -0.984 ± 0.000 | <0.0029 | Negative |
| Motor functions/Normal | -0.866 ± 0.173 | <0.0029 (except Birth) | Negative |
| Muscle Hypotonia | 0.223 ± 0.686 | <0.0029 (except Birth&5y) | Positive |
| NeuroDevelopmental Disorder-Seizures | -0.984 ± 0.000 | <0.0029 | Positive |
| QoL-Physical wellbeing (PW) | -0.984 ± 0.000 | <0.0029 | Negative |
| QoL-Psychological wellbeing (PsyW) | -0.984 ± 0.000 | <0.0029 | Negative |
| QoL-Social relationships (SR) | -0.984 ± 0.000 | <0.0029 | Negative |
| Seizure Threshold | -0.984 ± 0.000 | <0.0029 | Negative |
| Seizures Frequency | 0.984 ± 0.000 | <0.0029 | Positive |
| Sleep-Dysregulation/Loss | 0.984 ± 0.000 | <0.0029 | Positive |
| Speech & Language-Impaired | 0.984 ± 0.000 | <0.0029 | Positive |

**Table 8:**
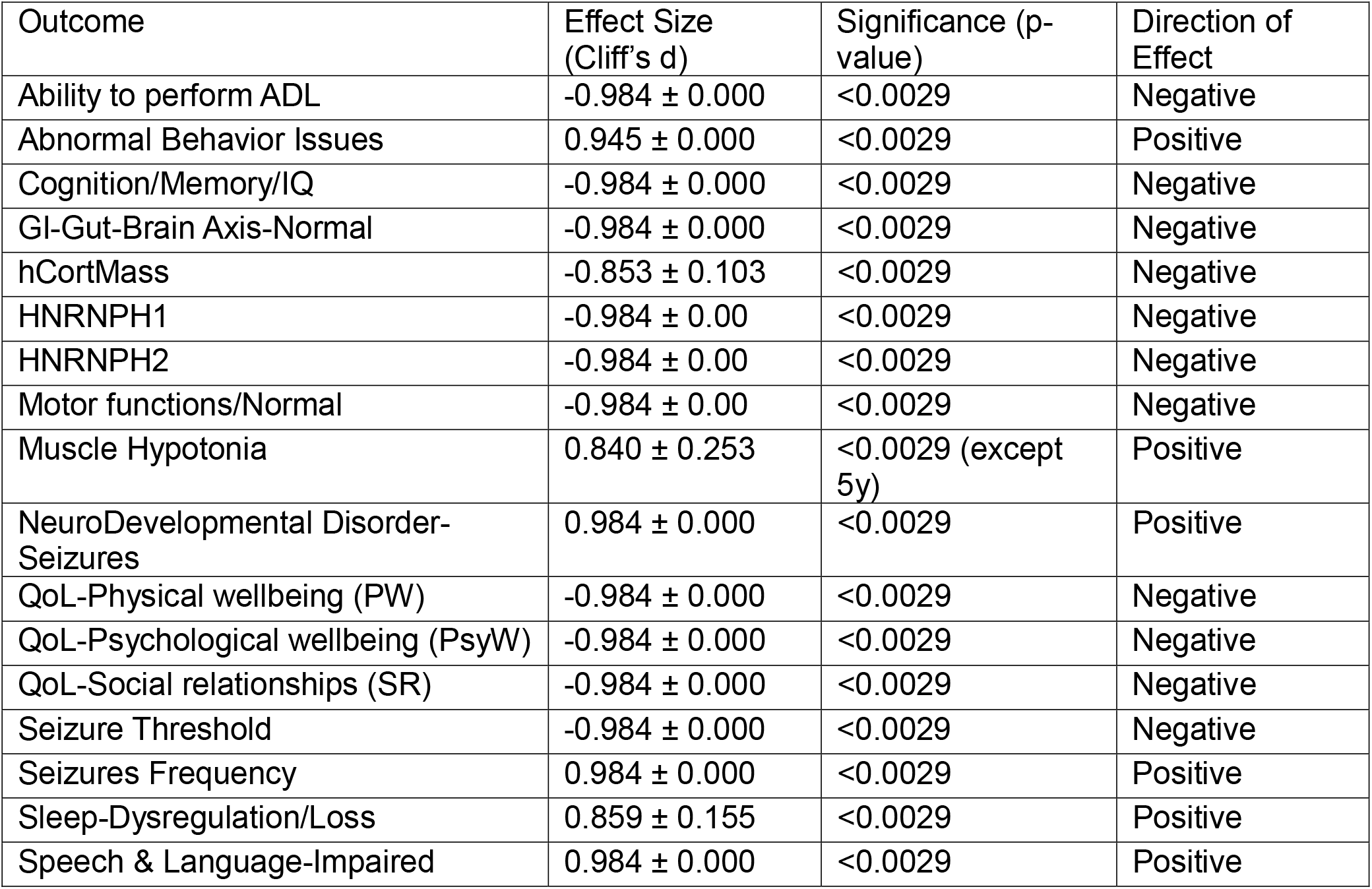
Effect Sizes and Significance Levels for comparison of NS/FSSevere vs. NS/FSMild HNRNPH2 LOF mutations.

|  |  |  |  |
| --- | --- | --- | --- |
| Outcome | Effect Size (Cliff's d) | Significance (p-value) | Direction of Effect |
| Ability to perform ADL | -0.984 ± 0.000 | <0.0029 | Negative |
| Abnormal Behavior Issues | 0.945 ± 0.000 | <0.0029 | Positive |
| Cognition/Memory/IQ | -0.984 ± 0.000 | <0.0029 | Negative |
| GI-Gut-Brain Axis-Normal | -0.984 ± 0.000 | <0.0029 | Negative |
| hCortMass | -0.853 ± 0.103 | <0.0029 | Negative |
| HNRNPH1 | -0.984 ± 0.00 | <0.0029 | Negative |
| HNRNPH2 | -0.984 ± 0.00 | <0.0029 | Negative |
| Motor functions/Normal | -0.984 ± 0.00 | <0.0029 | Negative |
| Muscle Hypotonia | 0.840 ± 0.253 | <0.0029 (except 5y) | Positive |
| NeuroDevelopmental Disorder-Seizures | 0.984 ± 0.000 | <0.0029 | Positive |
| QoL-Physical wellbeing (PW) | -0.984 ± 0.000 | <0.0029 | Negative |
| QoL-Psychological wellbeing (PsyW) | -0.984 ± 0.000 | <0.0029 | Negative |
| QoL-Social relationships (SR) | -0.984 ± 0.000 | <0.0029 | Negative |
| Seizure Threshold | -0.984 ± 0.000 | <0.0029 | Negative |
| Seizures Frequency | 0.984 ± 0.000 | <0.0029 | Positive |
| Sleep-Dysregulation/Loss | 0.859 ± 0.155 | <0.0029 | Positive |
| Speech & Language-Impaired | 0.984 ± 0.000 | <0.0029 | Positive |

